# Similar incidence of Coronavirus Disease 2019 (COVID-19) in patients with rheumatic diseases with and without hydroxychloroquine therapy

**DOI:** 10.1101/2020.05.16.20104141

**Authors:** Juan Macías, Paz González-Moreno, Esther Sánchez-García, Ramón Morillo-Verdugo, Carmen Domínguez-Quesada, Ana Pinilla, MªMar Macho, MªVictoria Martínez, Alejandro González-Serna, Anaïs Corma, Luis M. Real, Juan A. Pineda

## Abstract

**Background:** Hydroxychloroquine is currently being tested as post-exposure prophylaxis against coronavirus disease 2019 (COVID-19) in several ongoing clinical trials.

**Objective:** To compare the incidence of COVID-19 in Spanish patients with autoimmune rheumatic diseases treated with and without hydroxychloroquine.

**Methods:** Retrospective electronic record review, from February 27^th^ to April 16^th^, of patients with autoimmune inflammatory diseases followed at two academic tertiary care hospitals in Seville, Spain. The cumulative incidence of COVID-19, confirmed or suspected, was compared between patients with and without hydroxychloroquine as part of their treatment of autoimmune inflammatory diseases.

**Results:** Among 722 included subjects, 290 (40%) were receiving hydroxychloroquine.During the seven-week study period, five (1.7%[95% CI: 0.5%–4.0%] cases of COVID-19 were registered among patients with hydroxychloroquine and five (1.2%[0.4%2.7%])(p=0.523) in without hydroxychloroquine. COVID-19 was confirmed by PCR in one (0.3%, 95% CI 0.008-1.9%) patient with hydroxychloroquine and two(0.5%,95% CI 0.05%–1.6%) without hydroxychloroquine (p=1.0). One patient on hydroxychloroquine and two subjects without hydroxychloroquine were admitted to the hospital, none of them required to be transferred to the intensive care unit and no patient died during the episode.

**Conclusions:** The incidence and severity of COVID-19 among patients with autoimmune rheumatic diseases with and without hydroxychloroquine was not significantly different. Hydroxychloroquine does not seem to be an appropriate therapy for post-exposure prophylaxis against COVID-19.

## Introduction

Coronavirus disease 2019 (COVID-19) pandemic is currently a health emergency which has caused around 250.000 deaths worldwide in four months. Because of this, finding effective therapy and prophylactic strategies has become a maximum priority (1). Hydroxychloroquine (HCQ) has shown to be active against severe acute respiratory syndrome coronavirus 2 (SARS-CoV-2) *in vitro* by inhibiting several steps of the viral replication cycle, including some of the earliest ones, as the fusion to cell membrane (2). For this reason, and despite the *in vivo* effectivity of HCQ is still a matter of controversy (3,4), using this drug has been proposed as a rational strategy for post-exposure prophylaxis of SARS-CoV-2 infection (5). Indeed, at least nine clinical trials aimed to test this hypothesis are currently ongoing (6).

HCQ is commonly used as a part of the therapy of several autoimmune inflammatory diseases, such as rheumatoid arthritis (RA) or systemic lupus erythematous (SLE). If HCQ was effective as post-exposure prophylaxis of SARS-CoV-2, a reduced incidence of COVID-19 could be expected in patients receiving treatment with this drug. Data on this issue may provide us with worthy information on the potential of HCQ therapy as a prophylactic strategy, which may help to better design clinical trials. Because of this, in this study we aimed to compare the incidence of COVID-19 in Spanish patients with autoimmune rheumatic diseases treated with HCQ and without HCQ therapy, during seven weeks in the pandemic.

## Methods

This was a retrospective study, where subjects with autoimmune inflammatory disorders in which HCQ is commonly used, attending the Rheumatology Unit of Virgen Macarena University hospital and the Internal Medicine Unit of Valme University Hospital, both in Seville (Spain), were included. In these patients, we evaluated the cases diagnosed with COVID-19 from February 27^th^ to April 16^th^. In this period, 182816 confirmed COVID-19 cases were reported in Spain (7).

To evaluate the number of COVID-19 cases among the study patients, we searched for episodes of attendance to hospitals because of COVID-19 related symptoms in the shared electronic medical record of the hospitals of the Andalusian Health Service. COVID-19 was defined according to the Spanish Health Ministry criteria (8), as: 1) Confirmed: SARS-CoV-2 PCR in nasopharynx swab, serum antibody or antigen turned out to be positive. 2) Likely: serious acute respiratory infection, with symptoms and chest X-Ray signs consistent with COVID-19, but without microbiological confirmation. 3) Possible: mild acute respiratory infection during the study period, where microbiological studies were not carried out.

In the statistical analysis, continuous variables were compared by the Mann-Whitney U test and the categorical ones by the Fisher test. For the main rates, percentages and 95% confidence intervals (95% CI) were estimated. Statistical analyses were conducted using the package STATA 16.0 StataCorp, College Station, TX, USA.

The study was approved by the Ethic Committee of the Valme University Hospital.

## Results

Seven hundred and twenty-two subjects were studied. Two hundred and ninety (40%) of them were receiving HCQ. The main features of the study population, including those who were on HCQ therapy and those who were not, are shown in table 1.

During the seven-week study period, five (1.7% [95% CI: 0.5%–4.0%] cases of COVID-19 were registered among patients treated with HCQ and five (1.2% [0.4%–2.7%]) (p=0.523) in the group of subjects who were not receiving this drug. Most cases among both groups of patients met criteria for possible COVID-19 (four vs. two cases, respectively). Confirmed COVID-19 was diagnosed in one (0.3%, 95% CI 0.008–1.9%) subject treated with HCQ and two (0.5%, 95% CI 0.05%–1.6%) without this drug (p=1.0) (table 1). One patient on HCQ and two subjects without HCQ were admitted to the hospital, none of them required to be transferred to the intensive care unit (ICU) and no patient died during the episode.

**Table 1.** Features of thx study patients, according to they were on HCQ or not (n=722)

| Characteristic | HCQ<br>(n=290) | No HCQ<br>(n=432) | p value |
| --- | --- | --- | --- |
| Median (Q1-Q3) age, years | 56 (45-65) | 58 (48-68) | 0.140 |
| Male sex, n (%) | 42 (16) | 82 (21) | 0.123 |
| Rheumatic disease, n (%) |  |  | <0.001 |
| RA | 144 (50) | 323 (75) |  |
| SLE | 83 (30) | 11 (2.5) |  |
| Other | 60 (21) | 98 (23) |  |
| Immunosuppressive treatment,<br>n (%) | 169 (58) | 321 (74) | <0.001 |
| Anti-TNF drugs, n (%) | 3 (1) | 54 (13) | <0.001 |
| Latest median (Q1-Q3) CRP,<br>mg/dl before diagnosis | 2.8 (1.1-6.4) | 3.5 (1.4-8.2) | 0.046 |
| COVID-19 diagnosis, n (%) | 5 (1.7) | 5 (1.2) | 0.523 |
| Confirmed | 1 (20) | 2 (40) |  |
| Likely | 0 | 1 (20) |  |
| Possible | 4 (80) | 2 (40) |  |
| Admissions, n/N (%) | 1/5 (20) | 2/5 (40) | 1.0 |

HCQ: Hydroxychloroquine RA: Rheumatoid arthritis; SLE: Systemic lupus erythematosus;TNF: Tumor necrosis factor alpha. CRP: C-reactive protein. Other:Includes mixed connective tissue disease, scleroderma, spondylarthritis,undifferentiated polyarthritis, reactive arthritis, sarcoidosis, Sjogren syndrome, antiphospholipid syndrome, dermatomyositis, and Behçet disease.

Immunosuppressive treatment includes: abatacept, azathioprine, baricitinib, belimumab,corticosteroids, cyclophosphamide, leflunomide, methotrexate, mycophenolate,tacrolimus, tofacininib, anti-CD20, anti-IL1, anti-IL6, anti-IL12 and anti-IL23 and anti-IL17drugs.

## Discussion

According to these results, HCQ treatment does not seem to be an appropriate therapy for post-exposure prophylaxis against SARS-CoV-2. In fact, the incidence of COVID- 19 among patients with autoimmune rheumatic diseases on HCQ treatment was not significantly different of that observed in subjects not receiving HCQ. In addition, the severity of COVID-19, as measured by the rate of admission or ICU requirement, was similar in both groups.

Patients with immune-mediated inflammatory diseases are a population with particular features, that includes frequent use of biologic agents, other immunomodulatory drugs or both. This fact could increase the incidence or worsen the outcome of COVID-19. Howevere, preliminary data from a case-series in New York City suggested that the use of these drugs is not associated with a different clinical profile in COVID-19 (9). In addition, during the study period, in the province of Seville, an area with 1940000 inhabitants, 2278 confirmed COVID-19 cases were reported (10). This yields an overall incidence of 0.11% (95% CI 0.11%–0.12%) cases. This figure is inside the 95% CI of confirmed COVID-19 found herein both in subjects taking HCQ and in those who were not on this therapy.

This study has a few limitations. First, most cases were possible or likely COVID-19 cases, but shortage of diagnostic kits and health care system collapse in Spain and other countries have led to the fact than many cases of COVID-19 were not able to be confirmed. In addition, data come only from hospital registers and mild cases could have been seen only in primary care; likewise, most asymptomatic cases would have gone unnoticed. However, even having omitted patients, these data show that incidence of COVID-19 in subjects treated with HCQ would be far from zero. Therefore, HCQ would not be an ideal therapy for post-exposure prophylaxis of SARS-CoV-2 infection.

In summary, although the definitive information on the effectiveness of HCQ in post-exposure prophylaxis against SARS-CoV-2 infection will eventually come from controlled clinical trials, according to these results, other strategies should be designed to be tested, since HCQ monotherapy, as the best, does not protect against a significant number of infections with this coronavirus.

## Data Availability

Anonymized data could be accessed if requested

## Notes

### Competing Interest Statement

The authors have declared no competing interest.

### Clinical Trial

NA

### Funding Statement

Not funded

## References

1 Lai CC, Wang CY, Wang YH, Hsueh SC, Ko WC, Hsueh PR. Global epidemiology of coronavirus disease 2019: disease incidence, daily cumulative index, mortality, and their association with country healthcare resources and economic status. Int J Antimicrob Agents 2020;55:105946.

2 Yao X, Ye F, Zhang M, Cui C, Huang B, Niu P,et al. In Vitro Antiviral Activity and Projection of Optimized Dosing Design of Hydroxychloroquine for the Treatment of Severe Acute Respiratory Syndrome Coronavirus 2 (SARS-CoV- 2).Clin Infect Dis 2020;Mar 9.pii: ciaa23710.1093/cid/ciaa237.[Epub ahead of print].

3 Gautret P, Lagier JC, Parola P, Hoang VT Meddeb L, Sevestre J,et al. Clinical and microbiological effect of a combination of hydroxychloroquine and azithromycin in 80 COVID-19 patients with at least a six-day follow up: A pilot observational study. Travel Med Infect Dis 2020;Apr 11:101663.10.1016/j.tmaid.2020.101663.[Epub ahead of print]

4 Chowdhury MS, Rathod J, Gernsheimer J. A Rapid Sxstematic Review of Clinical Trials Utilizing Chloroquine and Hydroxychloroquine as a Treatment for COVID-19. Acad Emerg Med 2020; May 2. 10.1111/acem.14005. [Epub ahead of print]

5 Mitjà O, Clotet B. Use of antiviral drugs to reduce COVID-19: Lancet Glob Health 2020; March 19. Published online: https://doi.org//10.1016/52214-109X(20)30114-5

6 https://clinicaltrials.gov/ct2/results?cond=COVID19&term=hydroxychloroquine+post-exposure+prophylaxis+&cntry=&state=&city=&dist=. Accessed May 3rd, 2020

7 Red nacional de vigilancia epidemiolόgica. nforme sobre la situaciόn de COVID-19 en España Informe COVID-19 no 23. 16 de abril de 2020. https://www.isciii.es/QueHacemos/Servicios/VigilanciaSaludPublicaRENAVE/EnfermedadesTransmisibles/Documents/INFORMES/Informes%20COVID-19/Informe%20n°%2023.%20Situaciόn%20de%20COVID-19%20en%20España%20a%2016%20de%20abril%20de%202020.pdf

8> Ministerio de Sanidad.Gobierno de España.https://www.mscbs.gob.es/en/profesionales/saludPublica/ccayes/alertasActual/nCov-China/documentos/Procedimiento_COVID_19.pdf

9 Haberman R, Chen A, Castillo R, Adhikari S, Hundesman D Covid-19 in immune-mediated inflammatory diseases – Case series from New York. New Engl J Med 2020; April 29.Published online.10.1056/NEJMc2009567.

10 Junta de Andalucía. Consejería de Salud y Familias. Comunicado Coronavirus 17 de abril 2020. https://www.juntadeandalucia.es/organismos/saludyfamilias/actualidad/noticias/detalle/234667.html

